# Combined Impact of Prior SARS-CoV-2 Infection and Vaccination on Antibody Presence

**DOI:** 10.1101/2021.09.08.21263268

**Authors:** Jennifer A Shuford, Michael D Swartz, David L Lakey, Kimberly A Aguillard, Stephen J Pont, Melissa A Valerio-Shewmaker, Eric Boerwinkle

**Affiliations:** Texas Department of State Health Services, Austin, TX USA; The University of Texas Health Science Center at Houston, School of Public Health in Houston, Houston, TX, USA; University of Texas System, Austin, TX, USA; The University of Texas Health Science Center at Houston, School of Public Health in Brownsville, Brownsville, TX, USA

## Abstract

As COVID-19 continues to spread rapidly and vaccine uptake stagnates, questions remain about the amount of SARS-CoV-2 antibodies present in the population induced by either SARS-CoV-2 infection, by a COVID-19 vaccine, or both.

The TEXAS <u>C</u>oronavirus <u>A</u>ntibody <u>RE</u>sponse <u>S</u>urvey (CARES) is a statewide seroprevalence program which utilizes the Roche S-test to detect antibodies to the SARS-CoV-2 spike protein and the Roche N-test to detect antibodies to the SARS-CoV-2 nucleocapsid protein, to monitor the combined impact of prior infection and the COVID-19 vaccine. The current sample size having both S- and N-test data and reported vaccination status is 8,846.

Participants with prior infection (i.e. N+) and with either partial or full vaccination have the highest proportion of those showing the maximum value of the S-test (80.95% and 83.07%, respectively). Using a permutation test, there is no statistically significant difference between the median S-test value for those that have had prior infection and are partially vaccinated versus those that have had prior infection and are fully vaccinated. These groups both show significantly higher median amount compared to the other three groups: N+/not vaccinated, N-/partially vaccinated, and N-/fully vaccinated (all p-values < 0.0001).

Unvaccinated individuals with prior infection have one of the lowest median S-test values. For participants with previous SARS-CoV-2 infection and a COVID-19 vaccine, the median S-test value is high and is not statistically different between those who are partially vaccinated and those who are fully vaccinated.

## Introduction

With COVID-19 once again rapidly spreading across communities and populations, and vaccine coverage stagnating, questions remain about the amount of SARS-CoV-2 antibodies present in the population induced by either SARS-CoV-2 infection, by a COVID-19 vaccine, or both. Presence of SARS-CoV-2 antibodies may not directly translate into degree of immunity, due to the complex dynamics of the cellular and humoral immune response and lack of neutralizing information, but it does provide information about the relative degree to which different segments of the populations are producing antibodies in order to inform both clinical and public health recommendations and practice.

## Methods

The TEXAS <u>C</u>oronavirus <u>A</u>ntibody <u>RE</u>sponse <u>S</u>urvey (CARES) is a statewide seroprevalence program designed to assess the frequency of antibody positive individuals across Texas, a large and diverse population. The design of the Texas CARES program has been described previously,^1,2,3^ but briefly includes sampling from employees of various retail and industrial sectors, K-12 and university students and educators, and patients and staff at Federally Qualified Health Centers. After the launch of Texas CARES, an immunoassay for detection of antibodies to the SARS-CoV-2 spike protein (Roche S-test) became available and was added to the immunoassay for detection of antibodies to the SARS-CoV-2 nucleocapsid protein (Roche N-test), so that we are able to monitor the combined impact of prior infection and the COVID-19 vaccine. Both tests use whole blood and have a sensitivity and specificity exceeding 97%.^4,5^ Vaccine status was determined by questionnaire at the time of enrollment. The current sample size having both S- and N-test data and reported vaccination status is 8,846.

## Results

The value of the S-test across five groups is shown in Figure 1. The upper measurement limit of the S-test assay is 2500 U/ml. Those participants who have had prior infection (i.e. N+) and have been either partially or fully vaccinated have the highest proportion of those showing the maximum value of the S-test (80.95% and 83.07%, respectively). Using a permutation test, there is no statistically significant difference between the median S-test value for those that have had prior infection and are partially vaccinated versus those that have had prior infection and are fully vaccinated. These groups both show significantly higher median amount compared to the other three groups: N+/not vaccinated, N-/partially vaccinated, and N-/fully vaccinated (all p-values < 0.0001). Individuals in the N+/not vaccinated group with very high S-test values likely reflect individuals having received the vaccine after completion of the study survey, but before participating in the blood draw for assay of the antibodies.

**Figure 1.**
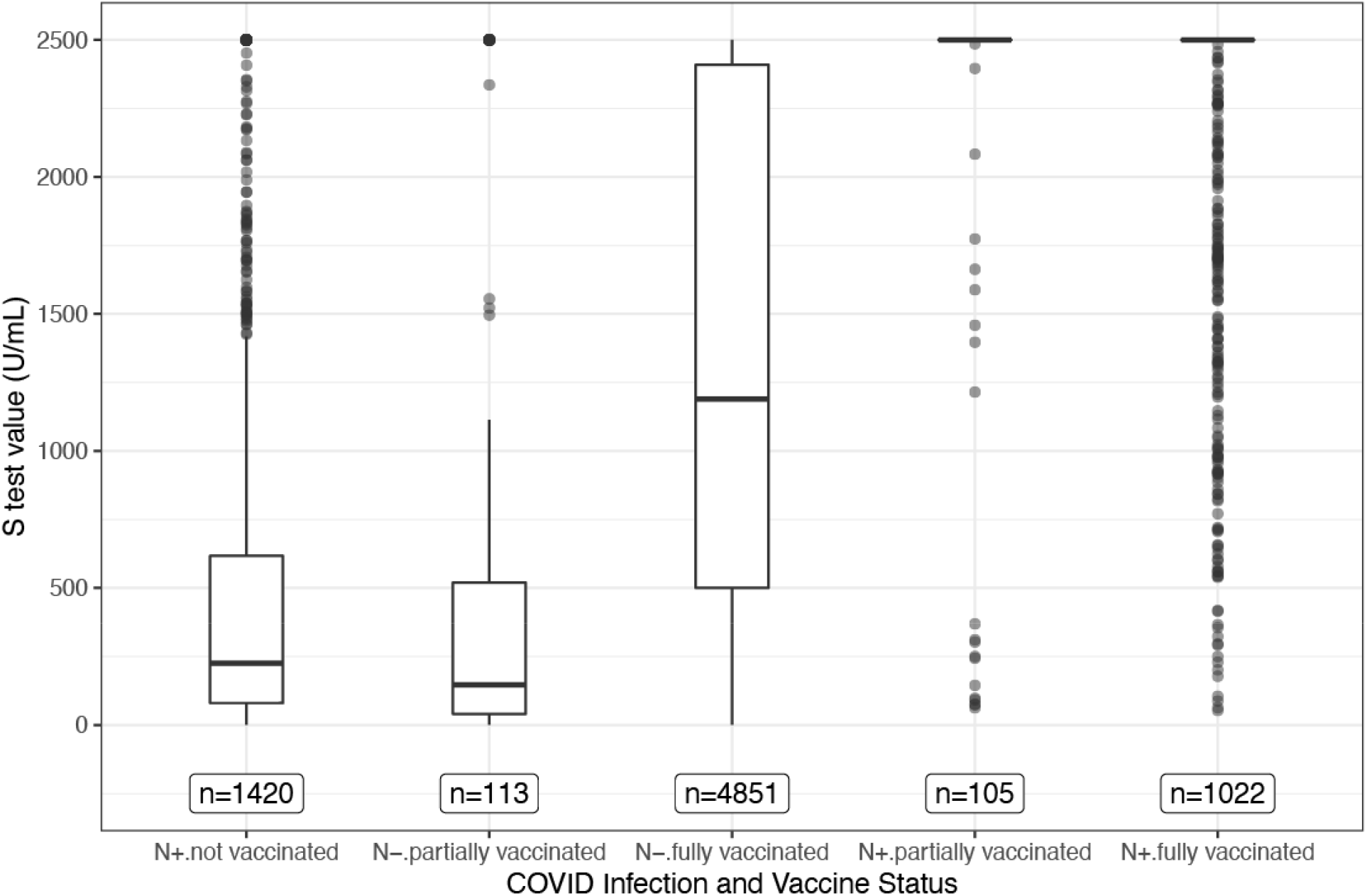
Boxplots of S-test values for each group. This figure shows a traditional boxplot for the S-test values for each group. The N-test status (+ or -) denotes prior infection (N+) or no prior infection (N-). Each box represents the data falling between the 25th and 75th percentiles. The horizontal bar within the box represents the median, and the whiskers extend 1.5 times the interquartile range below the 25th and above the 75th percentiles. The points that lie beyond the whiskers can be considered extreme values. Note that for the groups who had prior infection and either partial or full vaccination, over 80% of the values are greater than or equal to 2,500 U/mL, so the box part of the boxplot collapsed to the bar at the top of the graph.

## Discussion

The data reported here underscore two practical points. First, unvaccinated individuals with prior infection have one of the lowest median S-test values, perhaps indicating less protection than some have hoped for. Second, for those individuals participating in this project who were SARS-CoV-2 infected and then received the COVID-19 vaccine, the median S-test value is high and is not statistically different between those who are partially vaccinated and those who are fully vaccinated.

## Data Availability

Texas CARES investigators are committed to data sharing. Granular results and user-specified data summaries are currently publicly available on the Texas CARES portal. When baseline recruitment is complete, a deidentified individual level dataset will be available for download from the same portal.

https://sph.uth.edu/projects/texascares/dashboard

## Acknowledgements

This work was supported by the Texas Department of State Health Services and the University of Texas System. We would like to acknowledge the University of Texas Health Science Center at Houston, School of Public Health’s Texas CARES investigative team for their contribution to participant recruitment, data collection, statistical analysis, and data visualization including Sarah E Messiah, PhD; Melissa Valerio-Shewmaker, PhD, MPH; Steven Kelder, PhD, MPH; Harold W Kohl, PhD; Kimberly Aguillard, PhD; Michael Swartz, PhD; Stacia DeSantis, PhD; Ashraf Yaseen, PhD; Luis León-Novelo, PhD; Eric Boerwinkle, PhD; Jessica Ross, BS; Frances Brito, MS; Michael Gonzalez, MS; Leqing Wu, PhD; Onyinye OmegaNjemnobi, MBBS, MPH; Shiming Zhang, MS; Joy Yoo, BS; Tianyao Hao, MS; Cesar Pinzon Gomez, MD; Karina Farias, BA; Ashleigh Gil, MPH; David Lakey, MD; Jennifer Shuford, MD, MPH; Stephen Pont, MD, MPH.

This analysis would not have been possible without the partnership of many. The T CARES investigation team would like to thank Clinical Pathology Laboratories, Federally Qualified Health Care Centers statewide, and the Texas Association for Community Health Centers for assisting with sharing information with families about this survey.

